# Personalized circulating tumor DNA dynamics predict survival and response to immune checkpoint blockade in recurrent/metastatic head and neck cancer

**DOI:** 10.1101/2025.01.27.25321198

**Authors:** Daniel A. Ruiz-Torres, Ross D. Merkin, Michael Bryan, Julia Mendel, Vasileios Efthymiou, Thomas Roberts, Manisha Patel, Jong C. Park, Amber Chevalier, Clodagh Murray, Lisa Gates, Christodoulos Pipinikas, Shannon L. Stott, Adam S. Fisch, Lori J. Wirth, Daniel L. Faden

## Abstract

**Background:** Recurrent/metastatic head and neck squamous cell carcinoma (R/M HNSCC) is an aggressive cancer with a median overall survival of only 12 months. Existing biomarkers have limited ability to predict treatment response or survival, exposing many patients to the potential toxicity of treatment without certain clinical benefit. Circulating tumor DNA (ctDNA) has emerged as a non-invasive, real-time biomarker that could address these challenges.

**Methods:** We analyzed 137 plasma samples from 16 patients with R/M HNSCC undergoing immune checkpoint blockade (ICB)-based therapy. A tumor-informed, highly sensitive next-generation sequencing liquid biopsy assay (RaDaR, NeoGenomics Laboratories, Inc.) was applied to track ctDNA changes at baseline and throughout treatment. Univariable and multivariable analyses were used to assess the association between ctDNA negativity and key clinical outcomes: disease control (best objective response of stable disease, partial response, or complete response), three-year overall survival (OS), and three-year progression-free survival (PFS). We also assessed a machine learning model to predict disease progression based on ctDNA dynamics.

**Results:** Multivariable analysis revealed that ctDNA negativity during treatment was significantly associated with improved disease control (OR 21.7, 95% CI 1.86-754.88, p=0.0317), three-year OS (HR 0.04, 95% CI 0.00-0.47, p=0.0103), and three-year PFS (HR 0.03, 95% CI 0.00-0.37, p=0.0057). The machine learning model predicted disease progression with 88% accuracy (AUC 0.89).

**Conclusion:** Serial ctDNA monitoring predicted disease control, survival, and progression in patients with R/M HNSCC receiving treatment with ICB, suggesting that incorporation of ctDNA into clinical practice could enhance treatment decision-making for clinicians and improve patient outcomes.

## Introduction

Recurrent and/or metastatic head and neck squamous cell carcinoma (R/M HNSCC) is an aggressive cancer. Only a minority of patients benefit from the current standard-of-care options, including first-line therapies, which for most patients consist of immune checkpoint blockade (ICB) with or without concurrent cytotoxic chemotherapy. Even though objective response rates (ORR) are higher in PD-L1 expressing tumors (4.5% vs. 14.5% for pembrolizumab monotherapy and 31% vs. 34% for pembrolizumab plus chemotherapy in PD-L1 combined positive score (CPS) <1 vs 1-19 tumors), according to these data 66% to 95% of patients will not respond to ICB-based treatment (1–3). When compared with cetuximab-based regimens, ICB is associated with an improved median overall survival (OS) ranging from 12.3 to 14.9 months. However, the prognosis remains poor, highlighting a clear unmet need to improve therapeutic approaches for patients with R/M HNSCC (3). Despite its clear limitations as a predictive biomarker in R/M HNSCC, (4) PD-L1 is currently the only FDA-approved predictive biomarker to aid clinical decision-making in R/M HNSCC (1). Additionally, as a static biomarker (5), PD-L1 is only useful for predicting outcomes at the time of treatment initiation and cannot assist in clinical decision-making once a patient has started treatment. This underscores the need for dynamic, non-invasive biomarkers that can provide ongoing insights into disease status, enabling clinicians to tailor treatment decisions in real time.

Patients with R/M HNSCC that are resistant to ICB and have surgically salvageable disease who had either previously declined surgical approaches or, due to cytoreduction on immunotherapy, lesion(s) become amenable to surgical resection, may experience better local control with the appropriate application of salvage therapy (6,7). A non-invasive biomarker measured serially during therapy has the potential to augment the assessment of disease status and, thus, aid clinicians in personalizing the addition of local treatment modalities, such as surgery or radiotherapy, to systemic therapy, moving on to a new line of systemic therapy, and potentially providing a treatment holiday in the event of positive response to treatment.

Liquid biopsy with tumor-informed ctDNA offers an additional modality to monitor disease that is complementary to routine imaging methods currently used for assessing R/M HNSCC treatment response. This minimally invasive, real-time approach uses molecular methods to detect circulating tumor DNA (ctDNA) in clinically accessible biofluids, offering potential utility for treatment response assessment in the metastatic setting (8–12). ctDNA detection using next-generation sequencing (NGS) has strong predictive and prognostic potential across various solid tumors treated with ICB (13–15). Specifically, a decrease in ctDNA levels after one cycle of ICB has been associated with improved OS and progression-free survival (PFS) in multiple solid tumors (16). Importantly, ctDNA changes after one cycle of ICB have shown strong prognostic potential for both OS and PFS in HNSCC using tumor-naive and tumor-informed ctDNA detection methods (12,17). Similar results have been shown in patients with non-small cell lung cancer (NSCLC) (18–20), colorectal (16), urothelial (21), and HPV-associated tumors such as cervical (22) and anal squamous cell carcinoma (23).

Beyond estimating survival, ctDNA detection has also shown promising results in predicting disease progression in patients with HNSCC undergoing ICB. In a cohort of fourteen patients, tumor-informed ctDNA changes correlated with disease status, with decreasing ctDNA levels during treatment for responders and increasing levels for non-responders (17). However, with a limit of detection of 0.01% (measured in estimated variant allele frequency [%eVAF]), more sensitive approaches are needed to improve the reliability of the assay. These data underscore the potential for measurement of early on-treatment ctDNA dynamics to augment the estimation of anti-tumor response to ICB for patients with R/M HNSCC.

Because PD-L1 is both a weak predictive biomarker in HNSCC at the time of diagnosis and is not of utility in monitoring response to ICB, there is a critical need to develop and validate non-invasive and effective biomarkers. ctDNA detection approaches have shown promise but have been limited in their by sensitivity. We therefore elected to perform this prospective single-institution study in which we applied a tumor-informed, 10-fold more sensitive (Limit of Detection, LoD95: 0.0011%) next-generation sequencing liquid biopsy assay (24,25) to assess ctDNA dynamics in patients with R/M HNSCC as a biomarker of response to ICB-based therapy. We hypothesized that (1) ctDNA becoming undetectable during treatment would be associated with improved OS and PFS, while (2) persistent ctDNA detection would identify patients who will progress on ICB for R/M HNSCC.

## Methods

### Patient recruitment

Patients were eligible for this study if they started treatment for R/M HNSCC (oropharyngeal [OPC], oral cavity, nasopharyngeal [NPC], sinonasal, laryngeal, or hypopharyngeal anatomical primary sites, as well as unknown primaries) at Massachusetts General Hospital Cancer Center (MGHCC) using a regimen containing ICB between January 2021 and December 2022. Archival FFPE tumor tissue was reviewed and confirmed by a head and neck pathologist (A.S.F). Sixteen patients were enrolled in this study. This study was approved by the Dana-Farber/Harvard Cancer Center Institutional Review Board (DFCI 18-653), and all patients provided written informed consent.

### Clinical specimens

Serial blood samples were prospectively collected before, during, and after ICB treatment, between January 2021 and December 2022. Due to the inherent nature of clinical care, blood for ctDNA analysis was not collected on a fixed schedule across all patients. Blood samples were drawn in EDTA tubes, and plasma was obtained after two rounds of centrifugation (10 mins at 1600 RCF and 3000 RCF). Plasma and buffy coat were then aliquoted and stored at –80°C. Blood samples collected before and during treatment were available for prospective plasma ctDNA testing, while clinical data were annotated retrospectively following plasma testing completion. Objective response rate (ORR) was investigator-assessed using RECIST 1.1 or by retrospective review of physical examination data in clinical notes when imaging was unavailable (**Table S1**). Patients were classified according to the liquid biopsy response evaluation criteria in solid tumors (LB-RECIST) proposed by Gouda et al. (26). OS, PFS, and ORR outcomes were evaluated, and patients were followed for clinical outcomes until 8/16/2024.

### Tumor-informed ctDNA detection assay (RaDaR) workflow and QC metrics

Paraffin-embedded archival tissue sections from either tumor biopsies (n=2) or surgical resection specimens (n=14) were sent to NeoGenomics Laboratories, Inc. (Durham, NC) and processed for DNA extraction and whole exome sequencing (WES) (27). WES was performed to a median read depth of 306x (range: 112x – 607x) (**Figure 1A**). Personalized panels were successfully designed for all patients, capturing a median of 48 variants (range: 43-50) and released for plasma ctDNA testing with a median of 45 variants confirmed during panel QC (range: 19-49). Tumor-level key WES quality control metrics, the number of variants in each personalized panel, and those remaining after panel QC, as well as complete panel-specific variant information and their status in both tumor DNA and buffy coat control DNA, are provided in **Tables S2-S3**, respectively.

**Figure 1.**
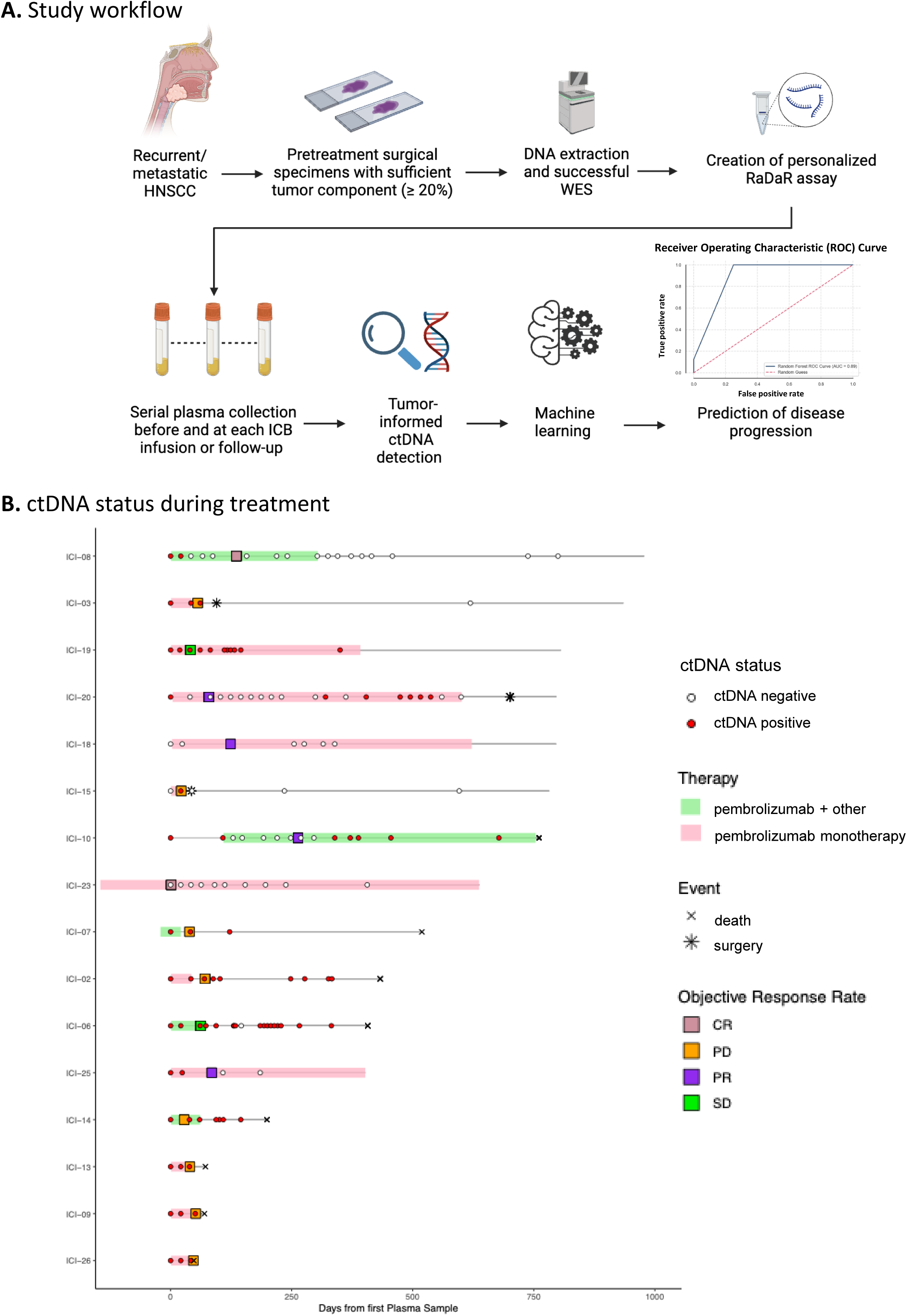
Tumor-informed ctDNA workflow and swimmer plot for the entire cohort. **A.** Archival tumor tissue from patients with head and neck squamous cell carcinoma undergoing ICB therapy was obtained and assessed. DNA was extracted for specimens with sufficient tumor components, and WES was performed to identify somatic mutations used to create personalized assays (RaDaR) for each patient. Plasma was obtained during ICB treatment, and each plasma sample was assessed for ctDNA detection. Longitudinal dynamics were used to feed the machine learning model, leading to an 88% accuracy in the prediction of disease progression. Figure created in BioRender.com. **B.** A swimmer plot shows longitudinal monitoring of ctDNA together with response assessed via clinical assessment or RECIST 1.1.

Cell-free DNA (cfDNA) was extracted from a median plasma volume of 3.6 mL (1.3 – 5.3 mL). Personalized panels were applied to serial plasma samples to assess ctDNA status before, during, and after treatment. Control genomic buffy coat DNA was included to identify and filter out confounding germline mutations and variants arising from clonal hematopoiesis of indeterminate potential (CHIP), thus eliminating false positive calls during plasma analysis. cfDNA and buffy coat control DNA were extracted at NeoGenomics Laboratories, Inc. (Durham, NC) using a solid-phase reverse immobilization magnetic bead protocol on a Hamilton Microlab STAR automated platform as previously described (27). ctDNA testing was performed using a median of 7,100 amplifiable copies (range: 1,000 – 20,000 copies) (**Table S4**) and a median depth of 233,350 reads per variant per sample (range: 82,597-740,848). The ctDNA status of all 137 samples tested across the 16 patients included in the study is shown in **Figure 1B**, alongside additional clinical information.

### Statistical analysis

This study assessed ctDNA negativity at any point during treatment as an independent predictor of treatment response and survival. To assess clinical and ctDNA predictors of best ORR per RECIST 1.1, we used univariable and multivariable logistic regression models to compute odds ratios (OR) with 95% confidence intervals (95% CI). To assess ctDNA as a pre-treatment or on-treatment predictor of disease progression (target outcome), we applied a machine learning model using three key features that we hypothesized would be associated with disease progression and could be obtained early in the treatment course: [1] detection of ctDNA before the start of treatment, [2] detection of ctDNA before imaging- or clinically-determined disease progression (but after the first infusion), [3] total increase or decrease in %eVAF from the baseline sample (P1) to the following sample (P2). A median of one cycle of ICB was administered in this timeframe (eleven patients received one cycle of ICB before the second ctDNA measurement, two patients received two cycles of ICB before the second ctDNA measurement, three patients were excluded from this analysis because either they had no pre-treatment samples or never had a sample positive for ctDNA). Patients were split 80/20 between training and testing, respectively. Using Stratified K-Fold cross-validation (5 folds), we ensured each fold mirrored the overall data distribution. We applied logistic regression within each fold, calculated key performance metrics— accuracy, precision, recall, F1-score, and area under the ROC curve (AUC)—and averaged these across iterations. Our model was reinforced by repeating the training and validation process 1000 times using bootstrapping, which tested the model against overfitting and underfitting.

To assess the association between ctDNA and both OS and PFS, Kaplan-Meier analysis was applied using the Log-rank test. We also performed univariable and multivariable Cox regressions and computed hazard ratios (HR) and 95% CI to assess the association between ctDNA negativity at any point during the study and OS and PFS. All values were considered statistically significant if α < 0.05. All analyses were performed using R Statistical Software (v4.3.1, R Core Team 2023), GraphPad (v10.1.2), or Python (v3.11.4). In R, the aod package (v1.3.3) was used to perform logistic regression, and the survival (v3.5.5) and survminer (v0.4.9) packages were used for survival analyses, which included both Kaplan-Meier analysis and Cox proportional hazard model analysis.

## Results

### Patient baseline characteristics

One hundred thirty-seven blood samples were collected between January 2021 and December 2022, with a median of 6.5 samples per patient (range: 3-20) from sixteen patients with R/M HNSCC being treated with ICB therapy. The median age was 62 (range: 52-91). Oral cavity was the most common anatomic subsite (8/16, 50%), and most patients received pembrolizumab monotherapy (11/16, 69%). Most patients had a PD-L1 CPS ≥ 20 (11/16, 69%). Among patients with virally-driven tumors, three patients had HPV-associated OPC, one patient had HPV-associated NPC, and one patient had EBV-associated NPC.

### Plasma ctDNA testing results

The overall ctDNA detection rate across all samples was 57% (78/137) (**Figure 1B**). The median %eVAF was 0.1399 (range 0.0007-7.0738) (**Table S4**). Patient baseline characteristics are summarized in **Table 1**. 88% (14/16) of patients had a pre-ICB plasma sample (considered the baseline sample for this study) available for analysis, with 12/14 (85.7%) being ctDNA positive (median %eVAF: 0.28, Range: 0.004-1.8). The two patients with negative baseline ctDNA both had regionally or distant metastatic oral cavity SCC with small volume disease – ICI-18 had a 4.1 x 5.5 x 5.7 cm cystic neck mass and ICI-15 had an enlarging 2.5 x 1.5 cm lung mass.

**Table 1.**
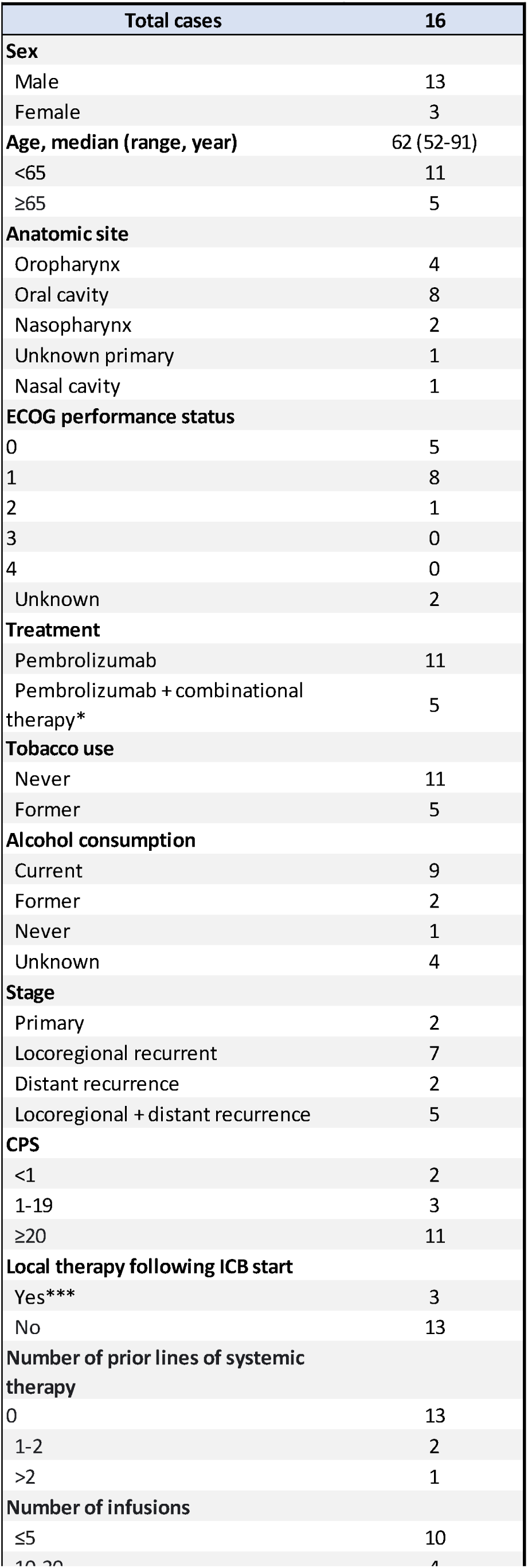
Clinical characteristics of the cohort. *Pembrolizumab + chemotherapy: n=4; Pembrolizumab + antiCD47: n=1; ***Salvage surgery: n=1; radiation n=2

The cohort consisted predominantly of patients belonging to LB-RECIST response groups 1 (detectable ctDNA that remains detectable after therapy; n=9) or 2 (detectable ctDNA that became undetectable after therapy; n=4). One patient was classified as Group 3 (undetectable ctDNA at baseline that becomes detectable after therapy), and two patients were classified as Group 4 (undetectable ctDNA that remains undetectable after therapy). Additionally, based on the quantitative response criteria categories, most patients showed ctDNA progressive disease (CPD; n=8) defined as an increase of >10% in VAF or de novo ctDNA detection or ctDNA complete response (n=4) defined as ctDNA clearance after initial detection (26,28) (**Table 2**). The median follow-up time was 19 months (range 1.5-42.6). There were two patients with best ORR of complete response (12%, CR), four patients with partial response (25%, PR), two patients with stable disease (12%, SD), and eight patients with progressive disease (50%, PD) (**Table 2**). The median time to achieve ctDNA negativity was 2.5 months (range 0.7-20.6) and the median time for ctDNA detection after clearance was 7 months (range 1.3-9.3). Three patients died from progressive disease soon after starting ICB therapy (ICI-9, ICI-13, ICI-26). For the remaining thirteen patients, a median of 1 (range 0 to 5) subsequent lines of treatment were received (including systemic therapy and salvage or palliative local therapy) following disease progression on ICB therapy. A median of 4 ICB infusions were administered (range 2-35), and a median of 13 months (range 1.3-27) elapsed from ICB initiation until death from any cause.

**Table 2.** LB-RECIST and radiological RECISTv1.1 in the cohort. Group 1: Patients with detectable ctDNA which remains detectable after therapy; Group 2: Patients with detectable ctDNA which became undetectable after therapy; Group 3: Patients with undetectable ctDNA which became detectable after therapy; Group 4: Patients with undetectable ctDNA which remained undetectable after therapy. D: Detectable, U: Undetectable (27). RECIST v1.1 was able to be applied in 14 cases. For 2 patients imaging was not available and clinical assessment was used. PD: progressive disease, SD: stable disease, PR: partial response, CR: complete response.

| **Table 2. LB-RECIST and radiological RECISTv1.1 in the cohort** |  |  |  |  |
| --- | --- | --- | --- | --- |
| **LB-RECIST Group** | **Definition** | **N (%) LB-RECIST** | **RECISTv1.1** | **N (%)** |
| 1 | D -> D | 9/16 (56) | PD | 8/16 (50) |
| 2 | D -> U | 4/16 (25) | SD | 2/16 (12) |
| 3 | U -> D | 1/16 (6) | PR | 4/16 (25) |
| 4 | U -> U | 2/16 (12) | CR | 2/16 (12) |

Eight patients (ICI-02, ICI-03, ICI-06, ICI-07, ICI-08, ICI-10, ICI-14, and ICI-15) had samples available after stopping ICB. Seven halted ICB due to PD, whereas one patient stopped ICB due to adverse events but showed no evidence of recurrence (ICI-08). Among those who developed PD, three (ICI-03, ICI-06, and ICI-15) underwent multimodality salvage therapy and demonstrated a decline in ctDNA to undetectable levels. By contrast, the remaining four patients with unsalvageable disease (ICI-02, ICI-07, ICI-10, and ICI-14) exhibited an increase in ctDNA levels immediately after stopping ICB. Notably, in the patient who had achieved a complete response (ICI-08), ctDNA levels remained undetectable following cessation of ICB. Lead time from ctDNA clearance to corresponding imaging response could not be accurately assessed because blood sample collection for ctDNA analysis and imaging assessment only coincided for a small selection of patients.

### ctDNA negativity predicts disease control

Among the four patients who became ctDNA negative during ICB therapy (LB-RECIST Group 2; ICI-08, ICI-10, ICI-20, ICI-25), one had CR (ICI-08), and three had PR (ICI-10, ICI-20, ICI-25) (**Figure S1, Table S1**). Two of those four patients (ICI-08, ICI-20) experienced immune-related adverse events (colitis and pneumonitis) after 1 and 1.5 years on ICB, respectively. Early on ICB therapy, the sensitivity and specificity of ctDNA negativity to predict an overall response (CR or PR) were 75% and 100%, respectively. Patients without pre-ICB sample available (n=2) and with persistently negative ctDNA levels (n=1) were also included in the analysis. Notably, in our small cohort, when patients with SD (n=2) were excluded, sensitivity and specificity for overall response were both 100%. ctDNA was assessed after a median of two infusions (range 1 to 7) and, on average, within 13 days (range -4 to 142) of radiological (n=14) or clinical (n=2) confirmation of RECIST 1.1 assessment. Interestingly, two patients (ICI-6, ICI-19) were classified as having stable disease (SD) despite a positive ctDNA result (**Table 3**).

**Table 3.** Radiographic and ctDNA molecular response. SD: stable disease, CR: Complete response, PR: partial response, PD: progressive disease. ctDNA results after 2 infusions (range 1 to 7) and within 13 days of RECIST assessment (range -4 to 142).

| **Table 3. Radiographic and ctDNA molecular response** |  |  |  |  |
| --- | --- | --- | --- | --- |
| **Best Objective Response** | **Molecular response** |  | **Sensitivity** | **Specificity** |
|  | **ctDNA-** | **ctDNA+** |  |  |
| SD/CR/PR | 6 | 2 | 100% | 75% |
| PD | 0 | 8 |  |  |

SD: stable disease, CR: Complete response, PR: partial response, PD: progressive c ctDNA results after 3.3 infusions (range 1 to 7) and within 13 days of RECIST assess

We performed logistic regression analyses to assess undetectable ctDNA as a predictor of treatment response ([1] ORR: CR/PR versus SD/PD; and [2] disease control: CR/PR/SD versus PD). In univariable analyses, we found that ICB regimen, PD-L1 CPS, or line of therapy were not predictive of ctDNA clearance. In univariable analysis, undetectable ctDNA at any time during the study period (either pre-treatment or on-treatment) was predictive of a 21-fold increase in odds of achieving disease control (CR, PR, or SD) (95% CI 2.04-558.55, p=0.0236). This association remained significant in multivariable analyses controlling for PD-L1, ICB regimen, and line of therapy (OR 21.7, 95% CI 1.86-754.88, p=0.0317). To ensure these results were not driven by the two patients in our cohort belonging to LB-RECIST Group 4 (patients who never had detectable ctDNA), we performed a sensitivity analysis by excluding ICI-18 and ICI-23 and observed that this finding persisted (univariable OR 15, 95% CI 1.37-408.3, p=0.04; multivariable OR 14.5, 95% CI 1.06-560.5, p=0.07).

### Undetectable ctDNA predicts improved overall and progression-free survival

Across all patients, the median OS was 19.9 months, and the median PFS was 2.7 months (**Figure 2 A-B**) from ICB initiation. In univariable analyses, we found no difference in OS or PFS when stratified by ICB regimen, PD-L1 CPS, recurrence type (locoregional, distant, or both), primary anatomical site, or virus (HPV or EBV) status. The absolute changes in ctDNA levels from pre-ICB to on-treatment samples were not associated with clinical outcomes. However, ctDNA negativity was found to be a significant predictor of improved OS and PFS. In univariable analyses, patients who had undetectable ctDNA at any point during the study period, including patients who did not have detectable ctDNA levels pre-ICB therapy (LB-RECIST Groups 2 and 4), had better OS (HR 0.10, 95% CI 0.02-0.48, p=0.001) and PFS (HR 0.17, 95% CI 0.04-0.69, p=0.006). In multivariable analyses controlling for PD-L1 CPS, ICB regimen, viral status, and recurrence type, the association remained significant for both OS (HR 0.04, 95% CI 0.00-0.47, p=0.0103) and PFS (HR 0.03, 95% CI 0.00-0.37, p=0.0057) (**Figure 2C-D**). We again performed a sensitivity analysis to ensure these results were not driven by patients who never had detectable ctDNA by excluding these two patients and found that the findings persisted (univariable HR for OS 0.17, 95% CI 0.03-0.86, p=0.0321; multivariable HR for OS 0.05, 95% CI 0-0.06, p=0.0175; univariable HR for PFS 0.23, 95% CI 0.06-0.94, p=0.0411; multivariable HR for PFS 0.05, 95% CI 0-0.61, p=0.019). ctDNA yielded comparable prognostic capability obtained with imaging-based prediction of OS. ORR (CR or PR) was associated with improved OS (HR 0.12, 95% CI 0.01-0.98, p=0.04) and disease control (CR, PR, or SD) trended toward improved OS (HR 0.28, 95% CI 0.07-1.64, p=0.08).

**Figure 2.**
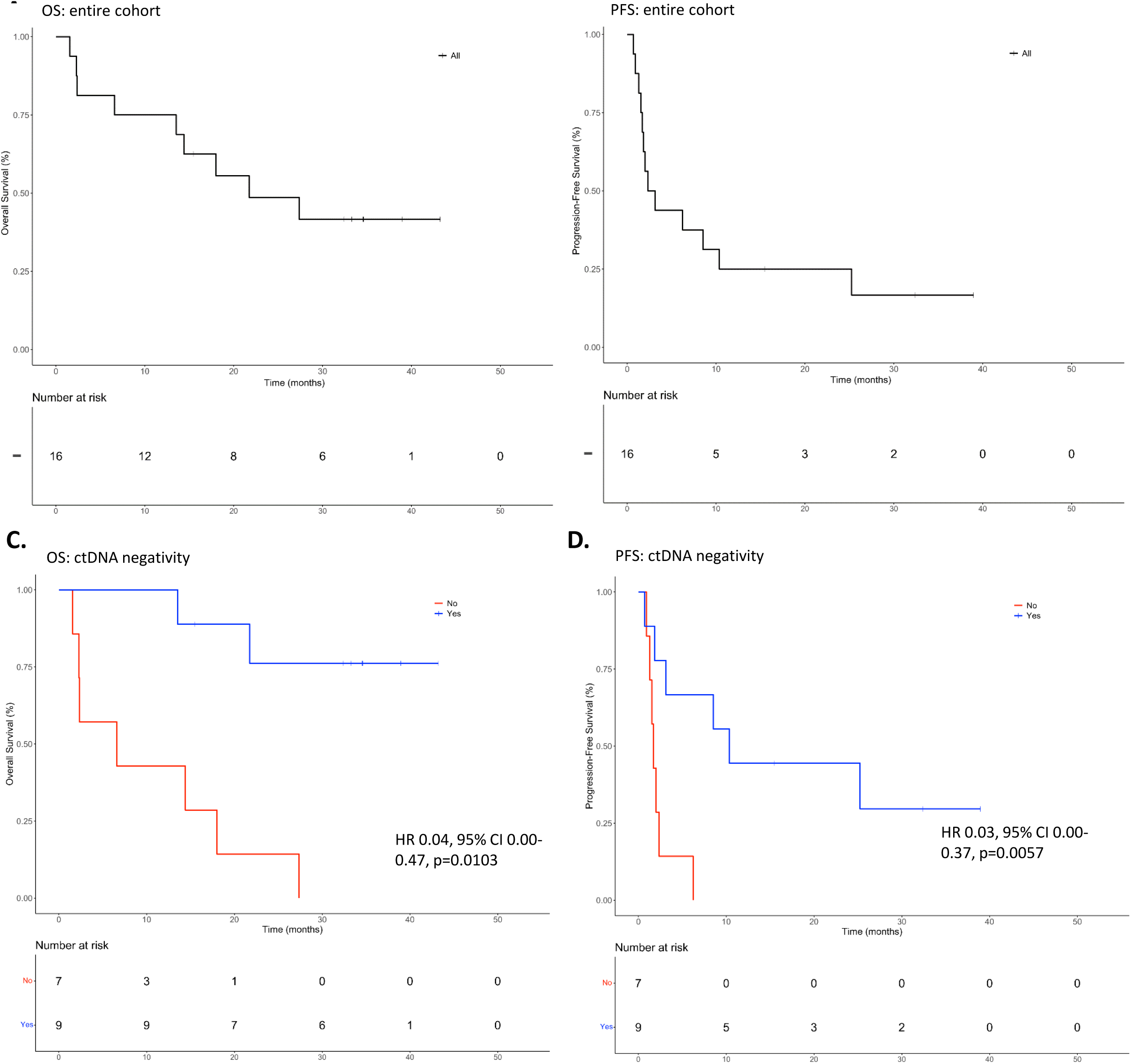
Three-year overall survival and progression-free survival for the entire cohort (A, B) and by ctDNA negativity at any point during the study period based on multivariable analysis (C, D).

### A machine learning model based on ctDNA dynamics predicts disease progression

We applied a machine learning model (random forest) using the following variables: [1] detection of ctDNA before the start of treatment, [2] detection of ctDNA before disease progression, and [3] total increase or decrease in %eVAF from the baseline sample (P1) to the following sample available (P2), to quantitatively assess ctDNA as a predictor of disease progression using metrics obtained early in the treatment course. We found the model was 88% accurate in correctly identifying patients who will experience disease progression (precision 80%, recall 100%, F1-score 89%, and area under the curve (AUC) 0.89), with a median lead time of 25 days (range 0-168) (**Figure 3A**). Feature importance analysis showed an increase in %eVAF early after a median of one cycle of ICB as the most impactful variable (**Figure 3B**). Notably, adding PD-L1 CPS to the model did not enhance its predictive performance.

**Figure 3.**
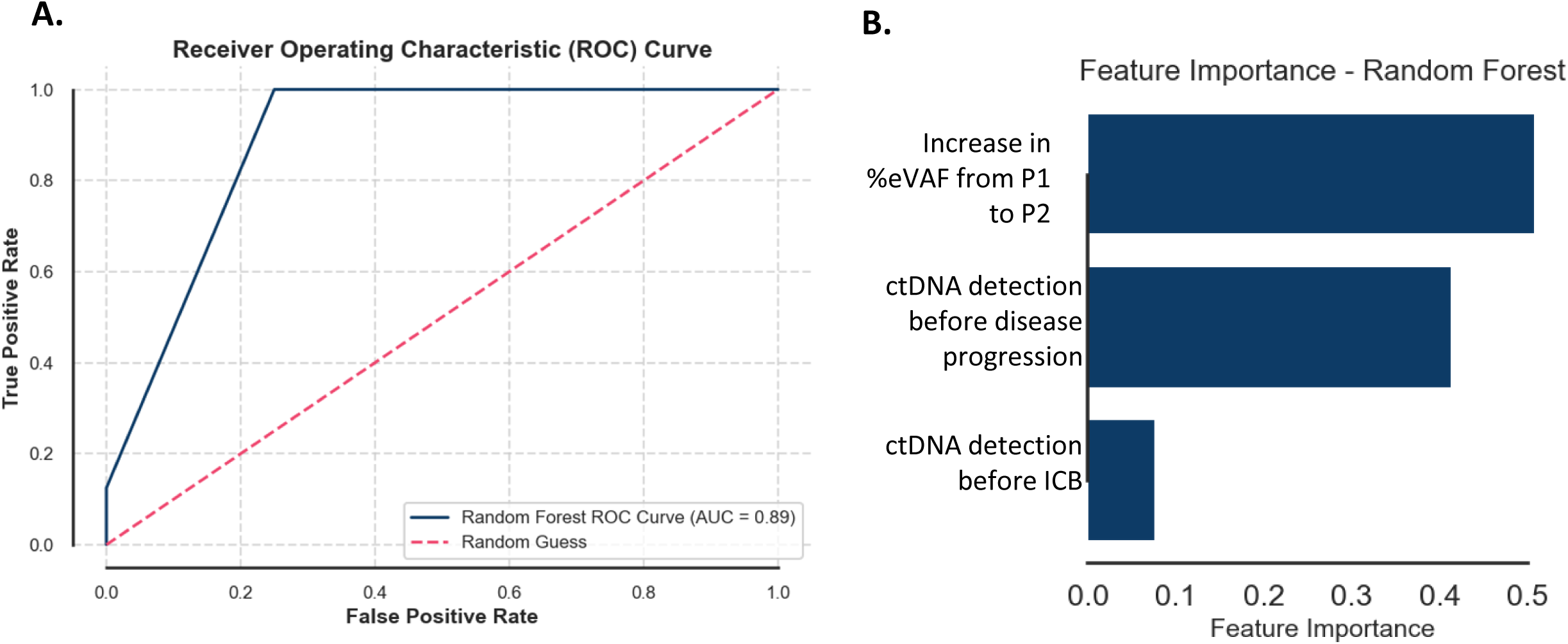
Area under the curve (A) and feature importance for the machine learning model (B).

## Discussion

This study evaluated the clinical utility of a tumor-informed WES-based liquid biopsy assay in predicting ORR, OS, PFS, and disease progression in 16 patients with R/M HNSCC receiving ICB-based therapy. Using serial ctDNA monitoring, we found ctDNA detection to be strongly predictive of disease progression, OS, and PFS, outperforming PD-L1 CPS, which is the only current FDA-approved predictive and prognostic biomarker in R/M HNSCC. In our cohort, patients became ctDNA negative through one of three ways: (1) patients who never had detectable ctDNA (LB-RECIST Group 4), (2) after ICB therapy (LB-RECIST Group 2), or (3) following salvage multimodality therapy (i.e., surgery or radiation) after progression on ICB (LB-RECIST Group 2). Having an undetectable ctDNA level conferred an overall and progression-free survival advantage irrespective of when the transition to becoming ctDNA undetectable occurred, including patients whose ctDNA was undetectable at baseline. We also found that, in patients who achieved a best response of CR, PR, or SD, serial ctDNA monitoring during treatment was 88% accurate in predicting disease progression after a median of 1 dose of ICB. . Taken together, these findings suggest that a tumor-informed liquid biopsy assay may aid clinicians in identifying patients with R/M HNSCC who could benefit from treatment escalation or next-line therapy.

Our study recapitulates findings reported in numerous other solid tumor types, including NSCLC, uveal melanoma, and microsatellite-unstable colorectal cancer (16,18,19,29). In these studies, the molecular response, as measured by serial ctDNA monitoring, was associated with radiographic response (29). For example, in a phase 2 clinical trial evaluating ctDNA in NSCLC, molecular response to ICB and chemotherapy showed a sensitivity of 82% and a specificity of 75% compared to RECIST 1.1 criteria as the gold standard (30). In contrast, our data indicate a lower sensitivity of 75% but a higher specificity of 100% in HNSCC, noting that these results should be interpreted in the context of a limited sample size. Importantly, in the NSCLC cohort, the molecular response was assessed after three cycles of ICB, and the median time to ctDNA negativity was 2.1 months, compared with a median of two doses of ICB and 2.5 months in our cohort, respectively.

Considering the limited sample size of our study, we wanted to verify that the association between ctDNA dynamics and ORR, OS, and PFS was not driven by the small subset of patients whose ctDNA was undetectable for the entire study period (LB-RECIST Group 4; ICI-18, ICI-23). In this sensitivity analysis, we re-ran all analyses excluding these two patients and observed similar results. The reason for undetectable ctDNA in these two patients is not clear but could be related to a low overall disease burden relative to other patients in the study or a low tumor proliferation rate (31). Additionally, one patient (ICI-23) had already received seven infusions before the first sample was available, potentially explaining persistent ctDNA negativity. Overall, these data align with prior reports that indicate having undetectable ctDNA at any time (LB-RECIST Groups 2 and 4) is associated with improved survival (16,27). Using the LB-RECIST classification system during our analysis helped to ensure a robust assessment of these data and highlights the utility of such a classification system to aid in a structured and systematic approach to analyzing ctDNA data sets. This is particularly important as ctDNA monitoring becomes even more commonplace. Harmonizing the collection and interpretation of ctDNA data across studies will strengthen the rationale for incorporating ctDNA monitoring into clinical practice across cancer types, disease settings, and treatment regimens.

Serial tumor-informed WES-based ctDNA monitoring has the potential to provide patients with more accurate prognostic information than the summary statistics typically reported by major clinical trials. This approach could also guide clinicians in tailoring subsequent treatment options for patients who fail to achieve undetectable ctDNA during ICB. Evidence suggests that escalation with multimodality interventions in the absence of molecular response can improve survival outcomes. For instance, although the SABR-COMET trial did not use ctDNA to determine eligibility, it demonstrated that stereotactic ablative radiotherapy in oligometastatic disease confers an overall survival advantage in multiple solid tumor types, including breast, lung, colorectal, and prostate (32). Incorporating serial ctDNA monitoring could serve as a valuable tool to better select patients with metastatic disease for local therapies, thereby optimizing treatment strategies and outcomes. A contrasting approach involves consideration of treatment de-escalation with a treatment holiday for patients with R/M disease who have sustained clearance of ctDNA on serial measurements. Our study demonstrated excellent survival in patients who clear their ctDNA and justifies considering this appealing de-escalation approach in prospective studies. Treatment holidays in this context could improve patient quality of life without compromising survival outcomes.

In addition to assessing the binary outcome of detectable/undetectable ctDNA, we also sought to characterize the predictive capability of ctDNA dynamics. We assessed various ctDNA dynamics parameters—absolute change in %eVAF after a median of one cycle, and thresholds of both >20% and >50% change in ctDNA levels from the pre-treatment to first on-treatment measurement (data not shown)—and found trends toward better outcomes in the early responders but no statistically significant findings. This is likely attributable to our small sample size and cohort heterogeneity and should be reassessed in a larger cohort of patients with R/M HNSCC. Our results support findings by Bratman and colleagues, who reported that a decrease in ctDNA levels after three cycles of pembrolizumab was predictive of achieving objective response (OR 28.74, 95% CI 3.51–253.04), independent of tumor type, tumor mutational burden, or PD-L1 level in a large cohort of patients with advanced solid tumors (n=94, HNSCC n=14) (15).

Among all patients with cancer regardless of tumor type, predicting in whom and when disease progression will occur represents a major unmet need. We therefore assessed ctDNA as a predictor of disease progression. Using a machine learning model, we leveraged on-treatment ctDNA dynamics, with a focus on changes in ctDNA early on treatment, to predict disease progression. Our findings are consistent with prior studies that indicate machine learning models can effectively predict response to immunotherapy in patients with solid tumors (33). In our cohort, the machine learning model was able to predict disease progression with a median lead time of 25 days (range 0-168). A lead time of approximately one month could minimize patient exposure time to ineffective treatment compared to the time it takes to identify progression by cross-sectional imaging. This additional time would also allow clinicians to explore and coordinate next-line therapies. Although ctDNA monitoring shows promise as a predictor of progressive disease, it remains an imperfect test. For one patient in this cohort (ICI-20), ctDNA was detected six days after imaging detection of PD, yet it was undetectable 15 days prior to imaging confirmation of PD. This finding underscores the importance of interpreting ctDNA results as complementary to, rather than a replacement for, evidence-based imaging approaches, thereby reducing the risks associated with false negative ctDNA results or the inability of even highly sensitive assays to detect rapidly evolving disease progression (26,31).

Our study has multiple limitations. First, a limited cohort size and the heterogeneity of the patient population (e.g., ICB regimen, a mixture of virus-associated and virus-independent tumors, PD-L1 CPS), limit the interpretation of our findings, although simultaneously improve their generalizability. Second, because samples were collected during routine clinical visits, our study did not apply a fixed schedule for sample collection, creating heterogeneity within the cohort, and this is particularly important in the context of lead time afforded by ctDNA in this study. This limitation should also be interpreted in the context that the optimal timing of serial ctDNA measurements for different systemic therapy regimens (e.g., immunotherapy versus chemoimmunotherapy) or tumor types is not known (31). Third, all clinical data were annotated retrospectively, possibly introducing retrospective bias. Fourth, although we applied a proprietary technology to characterize the serial ctDNA dynamics, the minimum clinically meaningful amount of ctDNA change between samples in patients with HNSCC has not yet been clearly defined (31).

In summary, our work supports further exploration of serial monitoring of ctDNA during treatment to predict disease control, PFS, OS, and progression in patients with R/M HNSCC receiving ICB-based therapy. Both treatment intensification in patients with persistently positive ctDNA on ICB therapy and de-intensification in patients with serially undetectable ctDNA on treatment should be further evaluated in prospective studies.

## Authorship contribution statement

D.L.F. conceptualized the investigation, provided supervision, and led the funding acquisition. Formal data analysis, including visualization, was performed by D.A.R.T., R.D.M., M.B.; D.A.R.T. and J.M. led the sample selection and shipping. J.M. and V.E. processed and stored the blood samples. A.S.F. identified the tumor on FFPE sections. D.A.R.T., R.D.M., T.R., M.P., J.C.P., C.P., L.J.W., and D.L.F. helped with review and editing. C.P., A.C., C.M., and L.G. processed WES samples and created the panel for variant detection in plasma samples. C.M. aided with data visualization. D.A.R.T. and R.D.M. wrote the original draft, and all authors performed critical reviews and revisions.

## Data availability statement

All the data relevant to this work is included in this manuscript.

## Funding

RaDaR assays were provided in kind for this study by NeoGenomics Laboratories, Inc, (Durham, NC).

## Declaration of interest

Daniel L. Faden receives research funding from Calico Life Sciences, in-kind funding from Boston Gene, Predicine, and NeoGenomics, receives consulting fees from Merck and Chrysalis Biomedical Advisors, and receives salary support from NIH 5K23DE029811, R03DE03550, 5R21CA267152. Shannon L. Stott receives research funding from NIH: 5R01CA226871-05, U18TR003793-02, V Foundation: T2020-004, American Cancer Society: 132030-RSG-18-108-01-TBG and MGH ECOR Scholars Program. Amber Chevalier, Christodoulos Pipinikas, and Clodagh Murray were NeoGenomics employees for the duration of the study.

## Supplementary Figures Legends

**Supplementary Figure 1.** ctDNA dynamics based on best objective response to ICB.

## Supplementary Tables Legends

**Supplementary Table 1**. Patient details. Group 1: Patients with detectable ctDNA which remains detectable after therapy; Group 2: Patients with detectable ctDNA which becomes undetectable after therapy; Group 3: Patients with undetectable ctDNA which becomes detectable after therapy; Group 4: Patients with undetectable ctDNA which remains undetectable after therapy. D: Detectable, U: Undetectable (27). CCR: ctDNA complete response -> ctDNA clearance after initial detectability. CPR: ctDNA partial response -> Decrease of >10% in variant allele frequency. CSD: ctDNA stable disease -> No increase or up to 10% increase or decrease in variant allele frequency. CPD: ctDNA progressive disease - > increase >10% in variant allele frequency or de novo ctDNA detection. CND: ctDNA non-measurable disease -> Undetectable ctDNA before and after treatment.

**Supplementary Table 2.** WES results. Patient ID: anonymized patient identifier; Panel ID: Identifier of personalized designed panel; Reads (million): Number of total sequencing reads resulting from whole exome sequencing; Aligned %: Percentage of reads correctly aligned to the reference genome; Duplication %: Percentage of duplicated reads; On target %: Percentage of reads falling within targeted regions (i.e. regions targeted by hybridization capture); Usable %: Percentage of usable reads (i.e., aligned, on target and not duplicated); Mean Coverage Tumor: mean read depth per nucleotide in Tumor, based on usable reads; Total Variants Panel: Number of variants included in personalized panel for RaDaR assay; Tumor Passing Variants: Number of variants that were confirmed for each patient by qualification testing of the patient panel; Mean tumor VAF QC passing variants: mean tumor VAF at panel QC for passing variants.

**Supplementary Table 3.** Variants per panel. Patient ID: Internal study patient identifier; Mutation code: Variant code summarizing location and nature of the variant included in the panel; Chromosome: Chromosome of the variant; Locus: Genomic locus of the variant (hg38 coordinates); Present in buffy coat: Boolean (TRUE/FALSE), indicating presence/absence in the buffy coat control (if FALSE the variant is retained for ctDNA assessment); VAF in WES: VAF (Variant Allele Frequency) in the Whole Exome Sequencing data; Plasma sample IDs: Semicolon-separated list of plasma sample identifiers; VAF in plasma: Semicolon-separated list of plasma VAFs, relative to their respective samples (Plasma sample IDs).

**Supplementary Table 4**. Plasma calling. Patient ID: Anonymized patient identifier; Panel ID: Identifier of personalized designed panel; Timepoint: Timepoint at which sample was collected; Collection Date: Date sample was collected; Specimen Type: Specimen Type; Accession ID: Order identifier of received patient samples; Sample ID: Sample identifier; Extracted Volume (ml): The volume of fluid used for extraction; Total Extracted Copies: The total amount of extracted DNA copies; Total Input Copies: The amount of DNA copies loaded into the RaDaR assay; eVAF%: The estimated mean variant allele fraction in the specimen, expressed as a percentage; Total Variants Panel: Total number of variants included in each personalized panel; Plasma Calling Variants: Number of variants passing all QC criteria per RaDaR panel and used in the final plasma ctDNA status (TRUE/FALSE) calling; Total Depth: Total plasma RaDaR sequencing depth per sample; Mean Depth: Mean plasma RaDaR sequencing depth per variant; ctDNA Detected: Information relating to whether circulating tumor DNA was detected or not (TRUE: Detected; FALSE: Not Detected).

## Notes

### Funding Statement

This study was funded by NIH 5K23DE029811; RaDaR assays were provided in kind for this study by NeoGenomics Laboratories, Inc. (Durham, NC).

### Author Declarations

Dana-Farber/Harvard Cancer Center Institutional Review Board gave ethical approval for this work

## References

1. Burtness B, Harrington KJ, Greil R, Soulières D, Tahara M, de Castro G, et al. Pembrolizumab alone or with chemotherapy versus cetuximab with chemotherapy for recurrent or metastatic squamous cell carcinoma of the head and neck (KEYNOTE-048): a randomised, open-label, phase 3 study. The Lancet. 2019 Nov;394(10212):1915–28.

2. Mehra R, Seiwert TY, Gupta S, Weiss J, Gluck I, Eder JP, et al. Efficacy and safety of pembrolizumab in recurrent/metastatic head and neck squamous cell carcinoma: pooled analyses after long-term follow-up in KEYNOTE-012. Br J Cancer. 2018 Jul 29;119(2):153–9.

3. Harrington KJ, Burtness B, Greil R, Soulières D, Tahara M, de Castro G, et al. Pembrolizumab With or Without Chemotherapy in Recurrent or Metastatic Head and Neck Squamous Cell Carcinoma: Updated Results of the Phase III KEYNOTE-048 Study. Journal of Clinical Oncology. 2023 Feb 1;41(4):790–802.

4. Burtness B, Rischin D, Greil R, Soulières D, Tahara M, de Castro Jr G, et al. Pembrolizumab Alone or With Chemotherapy for Recurrent/Metastatic Head and Neck Squamous Cell Carcinoma in KEYNOTE-048: Subgroup Analysis by Programmed Death Ligand-1 Combined Positive Score. Journal of Clinical Oncology. 2022 Jul 20;40(21):2321–32.

5. Lesterhuis WJ, Bosco A, Millward MJ, Small M, Nowak AK, Lake RA. Dynamic versus static biomarkers in cancer immune checkpoint blockade: unravelling complexity. Nat Rev Drug Discov. 2017 Apr 6;16(4):264–72.

6. Moon DH, Sher DJ. Oligometastasis in Head and Neck Squamous Cell Carcinoma. International Journal of Radiation Oncology*Biology*Physics. 2022 Nov;114(4):803–11.

7. Cohen EEW, Bell RB, Bifulco CB, Burtness B, Gillison ML, Harrington KJ, et al. The Society for Immunotherapy of Cancer consensus statement on immunotherapy for the treatment of squamous cell carcinoma of the head and neck (HNSCC). J Immunother Cancer. 2019 Dec 15;7(1):184.

8. Sivapalan L, Murray JC, Canzoniero JV, Landon B, Jackson J, Scott S, et al. Liquid biopsy approaches to capture tumor evolution and clinical outcomes during cancer immunotherapy. J Immunother Cancer. 2023 Jan 19;11(1):e005924.

9. Lapin M, Edland KH, Tjensvoll K, Oltedal S, Austdal M, Garresori H, et al. Comprehensive ctDNA Measurements Improve Prediction of Clinical Outcomes and Enable Dynamic Tracking of Disease Progression in Advanced Pancreatic Cancer. Clinical Cancer Research. 2023 Apr 3;29(7):1267–78.

10. Thompson JC, Scholes DG, Carpenter EL, Aggarwal C. Molecular response assessment using circulating tumor DNA (ctDNA) in advanced solid tumors. Br J Cancer. 2023 Dec 7;129(12):1893– 902.

11. Liu Z, Yu B, Su M, Yuan C, Liu C, Wang X, et al. Construction of a risk stratification model integrating ctDNA to predict response and survival in neoadjuvant-treated breast cancer. BMC Med. 2023 Dec 12;21(1):493.

12. Taylor K, Zou J, Magalhaes M, Oliva M, Spreafico A, Hansen AR, et al. Circulating tumour DNA kinetics in recurrent/metastatic head and neck squamous cell cancer patients. Eur J Cancer. 2023 Jul;188:29–38.

13. Zhang Q, Luo J, Wu S, Si H, Gao C, Xu W, et al. Prognostic and Predictive Impact of Circulating Tumor DNA in Patients with Advanced Cancers Treated with Immune Checkpoint Blockade. Cancer Discov. 2020 Dec 1;10(12):1842–53.

14. Lee JH, Menzies AM, Carlino MS, McEvoy AC, Sandhu S, Weppler AM, et al. Longitudinal Monitoring of ctDNA in Patients with Melanoma and Brain Metastases Treated with Immune Checkpoint Inhibitors. Clinical Cancer Research. 2020 Aug 1;26(15):4064–71.

15. Bratman S V., Yang SYC, Iafolla MAJ, Liu Z, Hansen AR, Bedard PL, et al. Personalized circulating tumor DNA analysis as a predictive biomarker in solid tumor patients treated with pembrolizumab. Nat Cancer. 2020 Aug 3;1(9):873–81.

16. Kansara M, Bhardwaj N, Thavaneswaran S, Xu C, Lee JK, Chang L, et al. Early circulating tumor DNA dynamics as a pan-tumor biomarker for long-term clinical outcome in patients treated with durvalumab and tremelimumab. Mol Oncol. 2023 Feb 13;17(2):298–311.

17. Hanna GJ, Dennis MJ, Scarfo N, Mullin MS, Sethi RKV, Sehgal K, et al. Personalized ctDNA for Monitoring Disease Status in Head and Neck Squamous Cell Carcinoma. Clinical Cancer Research. 2024 Jun 24;OF1–8.

18. Ricciuti B, Jones G, Severgnini M, Alessi J V, Recondo G, Lawrence M, et al. Early plasma circulating tumor DNA (ctDNA) changes predict response to first-line pembrolizumab-based therapy in non-small cell lung cancer (NSCLC). J Immunother Cancer. 2021 Mar;9(3):e001504.

19. Thompson JC, Carpenter EL, Silva BA, Rosenstein J, Chien AL, Quinn K, et al. Serial Monitoring of Circulating Tumor DNA by Next-Generation Gene Sequencing as a Biomarker of Response and Survival in Patients With Advanced NSCLC Receiving Pembrolizumab-Based Therapy. JCO Precis Oncol. 2021 Nov;(5):510–24.

20. Goldberg SB, Narayan A, Kole AJ, Decker RH, Teysir J, Carriero NJ, et al. Early Assessment of Lung Cancer Immunotherapy Response via Circulating Tumor DNA. Clinical Cancer Research. 2018 Apr 15;24(8):1872–80.

21. Powles T, Assaf ZJ, Davarpanah N, Banchereau R, Szabados BE, Yuen KC, et al. ctDNA guiding adjuvant immunotherapy in urothelial carcinoma. Nature. 2021 Jul 15;595(7867):432–7.

22. Kang Z, Stevanović S, Hinrichs CS, Cao L. Circulating Cell-free DNA for Metastatic Cervical Cancer Detection, Genotyping, and Monitoring. Clinical Cancer Research. 2017 Nov 15;23(22):6856–62.

23. Huffman BM, Singh H, Ali LR, Horick N, Wang SJ, Hoffman MT, et al. Biomarkers of pembrolizumab efficacy in advanced anal squamous cell carcinoma: analysis of a phase II clinical trial and a cohort of long-term responders. J Immunother Cancer. 2024 Jan 25;12(1):e008436.

24. Gale D, Lawson ARJ, Howarth K, Madi M, Durham B, Smalley S, et al. Development of a highly sensitive liquid biopsy platform to detect clinically-relevant cancer mutations at low allele fractions in cell-free DNA. PLoS One. 2018 Mar 16;13(3):e0194630.

25. Plagnol V, Woodhouse S, Howarth K, Lensing S, Smith M, Epstein M, et al. Analytical validation of a next generation sequencing liquid biopsy assay for high sensitivity broad molecular profiling. PLoS One. 2018 Mar 15;13(3):e0193802.

26. Gouda MA, Janku F, Wahida A, Buschhorn L, Schneeweiss A, Abdel Karim N, et al. Liquid Biopsy Response Evaluation Criteria in Solid Tumors (LB-RECIST). Ann Oncol. 2024 Mar;35(3):267–75.

27. Flach S, Howarth K, Hackinger S, Pipinikas C, Ellis P, McLay K, et al. Liquid BIOpsy for MiNimal RESidual DiSease Detection in Head and Neck Squamous Cell Carcinoma (LIONESS)—a personalised circulating tumour DNA analysis in head and neck squamous cell carcinoma. Br J Cancer. 2022 May 3;126(8):1186–95.

28. Gouda MA, Huang HJ, Piha-Paul SA, Call SG, Karp DD, Fu S, et al. Longitudinal Monitoring of Circulating Tumor DNA to Predict Treatment Outcomes in Advanced Cancers. JCO Precis Oncol. 2022 Jul;6:e2100512.

29. Cabel L, Riva F, Servois V, Livartowski A, Daniel C, Rampanou A, et al. Circulating tumor DNA changes for early monitoring of anti-PD1 immunotherapy: a proof-of-concept study. Annals of Oncology. 2017 Aug;28(8):1996–2001.

30. Anagnostou V, Ho C, Nicholas G, Juergens RA, Sacher A, Fung AS, et al. ctDNA response after pembrolizumab in non-small cell lung cancer: phase 2 adaptive trial results. Nat Med. 2023 Oct 9;29(10):2559–69.

31. Wyatt AW, Litiere S, Bidard FC, Cabel L, Dyrskjøt L, Karlovich CA, et al. Plasma ctDNA as a Treatment Response Biomarker in Metastatic Cancers: Evaluation by the RECIST Working Group. Clinical Cancer Research. 2024 Nov 15;30(22):5034–41.

32. Palma DA, Olson R, Harrow S, Gaede S, Louie A V., Haasbeek C, et al. Stereotactic Ablative Radiotherapy for the Comprehensive Treatment of Oligometastatic Cancers: Long-Term Results of the SABR-COMET Phase II Randomized Trial. Journal of Clinical Oncology. 2020 Sep 1;38(25):2830–8.

33. Li Y, Wu X, Fang D, Luo Y. Informing immunotherapy with multi-omics driven machine learning. NPJ Digit Med. 2024 Mar 14;7(1):67.

